# Serum NfL in spinocerebellar ataxia type 1 is increased already at the preataxic stage, correlating with proximity to clinical onset

**DOI:** 10.1101/2021.09.14.21263261

**Authors:** Carlo Wilke, David Mengel, Ludger Schöls, Holger Hengel, Maria Rakowicz, Thomas Klockgether, Alexandra Durr, Alessandro Filla, Bela Melegh, Rebecca Schüle, Kathrin Reetz, Heike Jacobi, Matthis Synofzik

## Abstract

**Background and Objectives:** Neurofilament light (NfL) appears a promising fluid biomarker in repeat-expansion spinocerebellar ataxias (SCAs), with piloting studies in mixed SCA cohorts suggesting that NfL might be increased at the ataxic stage of spinocerebellar ataxia type 1 (SCA1). We here hypothesised that NfL is increased not only at the ataxic stage of SCA1, but also at its – likely most treatment-relevant – preataxic stage.

**Methods:** We assessed serum (sNfL) and cerebrospinal fluid (cNfL) levels of NfL in both preataxic and ataxic SCA1, leveraging a multicentric cohort of 40 SCA1 carriers (23 preataxic, 17 ataxic) and >80 controls, and clinical follow-up data including actually observed (rather than only predicted) conversion to the ataxic stage (11 carriers).

**Results:** sNfL levels were increased with high age-corrected effect sizes at the preataxic (r=0.62) and ataxic stage (r=0.63), paralleling increases of cNfL levels. In preataxic subjects, sNfL levels increased with proximity to *predicted* ataxia onset, with significant sNfL elevations already 5 years before onset, and confirmed in preataxic subjects with *actually observed* ataxia onset. sNfL increases were detected already in preataxic SCA1 subjects without volumetric atrophy of cerebellum or pons, suggesting that sNfL might be more sensitive to early preataxic neurodegeneration than the currently known most change-sensitive regions in volumetric MRI. Using longitudinal sNfL measurements, we estimated sample sizes for clinical trials using the reduction of sNfL as endpoint.

**Conclusions:** sNfL levels might thus provide easily accessible peripheral biomarkers in both preataxic and ataxic SCA1, allowing stratification of preataxic subjects regarding proximity-to-onset, early detection of neurodegeneration even before volumetric MRI alterations, and potentially capture of treatment response in clinical trials.

## Introduction

Spinocerebellar ataxia type 1 (SCA1) is a devastating neurodegenerative disease characterised by rapid and irreversible decline of motor function already in mid-life, caused by a translated CAG repeat expansion in the *ATXN1* gene ^1-3^. The preataxic stage of SCA1 might provide a unique opportunity to delay or even prevent the neurodegenerative process by early therapeutic intervention, with targeted molecular therapies now coming into reach ^3^. Particularly, interventions with antisense oligonucleotides (ASOs) targeting mutated *ATXN1* have shown first promising results in mitigating the molecular, pathological, and behavioural phenotype in SCA1 mouse models ^4, 5^. Such ASO interventions might allow preventing the neurodegenerative process even before the occurrence of clinical symptoms ^6, 7^. However, to pave the way for upcoming clinical trials of these therapies, objective and easily accessible biomarkers are needed for both the preataxic and ataxic stage of SCA1, particularly for stratification of preataxic subjects in proximity to future clinical onset, early detection of neuronal decay, and capture of the treatment response. We here propose serum levels of neurofilament light (sNfL) as objective and easily accessible blood biomarkers for preataxic disease stratification and detection of early neuronal decay in SCA1. Neurofilaments are neuron-specific cytoskeletal proteins, released upon neuronal damage and, with ultra-sensitive assays, also reliably quantifiable in peripheral blood ^8-10^. Previous monocentric studies in mixed cohorts of repeat-expansion spinocerebellar ataxias (SCAs), including small subsets of SCA1 patients ^11, 12^, indicated that blood levels of NfL in multisystemic repeat-expansion SCAs are increased at the ataxic disease stage. However, these studies lacked detailed cohort-based assessment of the preataxic and conversion stage of SCA1, validation of blood NfL levels by cerebrospinal fluid (CSF) measurements of NfL (cNfL), and estimation of sample sizes for treatment trials using NfL as outcome parameter.

Leveraging a multicentric cohort of both preataxic and ataxic SCA1 subjects with longitudinal follow-up assessments – including not only the predicted, but also the actually observed ataxia onset – we here tested the hypothesis that blood levels of NfL in SCA1 may serve as a peripheral biomarker of (1) proximity to clinical ataxia onset, (2) early neuronal decay at the preataxic stage, as well as (3) objective trial outcome parameter allowing reduction of the required sample sizes in treatment trials. We expected increased serum and CSF levels of NfL already at the preataxic stage – with the sNfL increase in preataxic subjects preceding first signs of brain atrophy in volumetric MRI and rising further in proximity to ataxia onset – and even further NfL increase at the ataxic disease stage.

## Methods

### Cohort

Our multicentric cohort comprised 40 SCA1 subjects and 89 healthy controls, recruited within the framework of the EUROSCA (European integrated project on spinocerebellar ataxias) and RISCA (Prospective study of individuals at risk for spinocerebellar ataxia) consortia (recruitment period: 2008-2016, clinicaltrials.gov: NCT01037777) and additionally by the Department of Neurodegenerative Diseases, Center for Neurology, University of Tübingen (EUROSCA: 8 ataxic carriers; RISCA: 19 preataxic carriers, 19 controls; Tübingen: 4 preataxic carriers, 9 ataxic carriers, 70 controls) ^13-15^. The recruitment included subjects with genetically confirmed SCA1 (*ATXN1* repeat length ≥ 39), their first-degree relatives (i.e., siblings and children), and unrelated neurologically healthy controls. Carrier status was determined in all recruited relatives of mutation carriers. Based on their score on the Scale for the Assessment and Rating of Ataxia (SARA) ^16^, SCA1 mutation carriers were classified as either ataxic (SARA score ≥3, 17 subjects) or preataxic (SARA score <3, 23 subjects). Controls comprised mutation-negative first-degree relatives of SCA3 carriers and unrelated healthy individuals, all without symptoms or signs of neurodegenerative disease. Sample size calculation was based on a piloting study indicating that 8 ataxic SCA3 subjects and 8 controls would suffice to detect significant differences of sNfL levels between groups (assuming α=0.01, β=0.01, equal group size, two-tailed non-parametric test) ^12^. However, we included all available SCA1 subjects to also study associations of sNfL levels with clinical and genetic variables. Longitudinal blood samples were assessed in 17 subjects (11 ataxic, 3 preataxic, 3 control subjects; sampling interval: 2.7 years (2.0-3.4), median (IQR)). CSF levels of NfL (cNfL) were available of another 6 SCA1 subjects (5 ataxic and 1 preataxic subject, see Supplement 1 for details) and compared to cNfL levels of an independent cohort of 89 neurologically healthy controls (also recruited at the Department of Neurodegenerative Diseases, University of Tübingen). The repeat length of the expanded and normal alleles was determined by PCR-based fragment length analysis from 100–250 ng genomic DNA (CEQ8000 capillary sequencer, Beckman Coulter). Demographic, clinical, and genetic characteristics of both cohorts are summarised in Table 1.

**Table 1.** Baseline characteristics of the cohort with serum NfL measurements.

| group | preataxic carriers | ataxic carriers | controls |
| --- | --- | --- | --- |
| n | 23 | 17 | 89 |
| subjects with longitudinal samples | 3 | 11 | 3 |
| male sex | 26% | 53% | 45% |
| age (years) | 25.5 (22.4-32.8) | 44.7 (37.1-50.4) | 30.8 (27.1-40.5) |
| SARA score | 0.5 (0.0-1.0) | 12.5 (10.5-16.5) |  |
| repeat count | 47 (46-50) | 48 (44-51) |  |
| disease onset (years) |  | 35 (30.5-42.0) |  |
| disease duration (years) |  | 7.5 (4.9-9.0) |  |
| sNfL (pg/ml) | 15.5 (10.5-21.1) | 31.6 (26.2-37.7) | 6 (4.7-8.6) |

The table refers to the cohort for which serum measurements were done (see Supplement 1 for an analogous table on the characteristics of the cohort for which CSF measurements were done). SCA1 mutation carriers and controls did not differ significantly in sex (OR=1.36, $p=0.449$ , Fisher's exact test) and age ( $p=0.916$ , $U=2621$ , $z=0.11$ , $r=0.01$ , Mann-Whitney U-test). Data are reported as median and interquartile range. SARA: Scale for the assessment and rating of ataxia.

### Standard Protocol Approvals, Registrations, and Patient Consents

The ethics committee of the University of Tübingen approved the study (AZ 103/2017BO2). All subjects provided written informed consent before participation according to the Declaration of Helsinki.

### NfL quantification

Blood samples were centrifuged (4,000 *g*, 10 min, at room temperature). Serum was frozen at −80 °C within 60 min after collection, shipped and analysed without any previous thaw-freeze cycle. We measured sNfL levels in technical duplicates by single molecule array (Simoa) technique on the Simoa HD-X analyser (Quanterix, Billerica), using the NF-light Advantage kit according to the manufacturer’s instructions and reagents from a single lot (#502554) (dilution factor 1 in 4 in sample buffer) ^9^. Serum was centrifuged at 14,000 x *g* for 4 minutes and the upper 90% transferred to the assay plates. All measurements had a coefficient of variation (CV) below 20% (mean CV: 4.6%). All concentrations were in the previously established range of quantification ^9^, with high stability of the assay (within-run CV < 5.0%, between-run CVs < 1.2%). The lower limit of quantification (LLoQ) was 0.5 pg/ml across all runs, defined as the lowest standard having a signal higher than the average signal of the blank plus 9 SDs and allowing a recovery of in the range of 100% ± 20%. Longitudinal samples were measured in the same batch. We measured cNfL levels in technical duplicates by ELISA technique, using the UmanDiagnostics NF-light kit (Umeå, Sweden) according to the manufacturer’s instructions (dilution factor 1 in 2 in sample buffer, within-run CV < 5%, between-run CV < 10%, LLoQ 81 pg/ml) ^17^. Technicians were blinded to the genotypic and phenotypic status of the samples.

### Brain imaging

For quantification of pontine and cerebellar atrophy, which have been shown to represent the most change-sensitive volumetric MRI regions in SCA1 ^18, 19^, regional brain volumes were assessed in 12 preataxic carriers by semi-automated segmentation methods, based on T1-weighted volumetric MRI scans, as previously described ^15^. Signs of pontine or cerebellar atrophy were considered to be present if the respective brain volumes were below a threshold defined as the lower boundary of the 90% confidence interval (i.e., the 5% quantile) of the respective volumes measured in healthy age-matched controls (Supplement 2).

### Statistical analysis

#### Group effects

We used Mann-Whitney U-tests (two-sided, significance level: p<0.05, Bonferroni-corrected for multiple comparisons) to compare NfL baseline levels between phenotypic groups (i.e., preataxic, ataxic and control subjects). To correct the group effects for the age-related NfL increase observed in controls ^20, 21^, we calculated the z-score of each subject in relation to the NfL distribution in controls at the same age ^22^. For this, the difference between the measured level and the level predicted for controls at the same age was standardised relative to the distribution in controls at this age. Levels in controls were modelled by linear regression on the level of log-transformed data. Throughout the analyses, if the assumption of normality was violated (assessed by inspection and Shapiro-Wilk test), we used log-transformed data for the statistical analysis after ensuring that the transformed data were normally distributed. If normality was violated also after transformation, non-parametric tests were applied. We reported the effect size r of the applied tests wherever possible. We analysed the data in R (version 4.1).

#### Association of sNfL levels with age and repeat length

We analysed the association of sNfL levels with age and CAG repeat length in SCA1 carriers by linear regression. Specifically, we modelled sNfL levels (log-transformed) in all SCA1 carriers (n=40) with the predictors age and *ATXN1* CAG repeat length. We centred age at 34 years (i.e., mean age of carriers) and CAG repeat length at 47 repeats (i.e., mean CAG repeat length), as in previous analogous analyses ^22, 23^. We excluded one preataxic outlier (sNfL < 5 pg/ml) to fulfil model assumptions of the regression model.

#### Association of sNfL with proximity to ataxia onset

We analysed sNfL levels in all preataxic SCA1 subjects as a function of the time to the *predicted* onset of ataxia and, for the subset of 11 preataxic subjects converting to the ataxic stage during follow-up, as a function of the time to the *actually* observed onset of ataxia. For each preataxic SCA1 subject, we individually calculated the time to the predicted onset of ataxia based on the CAG repeat count and the age at the time of assessment, as established previously ^24^. Hereby the estimate based on the repeat size is adjusted for the age which the individual has actually reached without developing ataxia, meaning that the older the preataxic subject at the time of assessment, the higher the predicted age at onset. Moreover, we assessed the conversion-free follow-up duration of preataxic subjects by Kaplan-Meier analyses (analogous to the event-free survival times in clinical trials, R packages: *survival* and *survminer*). To test the hypothesis that baseline sNfL levels allow stratifying preataxic subjects regarding their time to phenoconversion, we used log-rank tests to compare the median time to conversion between preataxic subjects with high vs. low sNfL levels (threshold defined by median split).

#### Sample size estimation for intervention trials

We estimated the sample sizes required in future treatment trials using the reduction of sNfL levels towards levels observed in healthy controls as outcome measure ^22, 25^. We estimated the total sample size required to detect a given control-adjusted relative reduction of sNfL levels (20-80%) in the treatment arm, assuming that null mean change over time occurred in the placebo arm of the trial. We based the assumed inter-subject variability in the hypothetical trial on the measured intra-subject variability in the change of analyte levels (from baseline to first follow-up) in our ataxic subjects. The estimation further assumed equal numbers of subjects in both study arms (i.e., allocation ratio 1:1), α=0.01, β=0.01, two-tailed independent t-tests, and the use of log-transformed biomarker levels. It was performed with GPower 3.1 software (Kiel, Germany).

### Data availability

The anonymised data of this article can be accessed on reasonable request addressed to the corresponding authors.

## Results

### Serum NfL levels are increased already at the preataxic stage of SCA1, with further increases at the ataxic stage

sNfL levels were significantly increased not only in ataxic SCA1 subjects (31.6 pg/ml (26.2-37.7), median and interquartile range) (U=1666, z=6.51, p<0.001, Bonferroni-corrected for three groups), but also in preataxic SCA1 subjects (15.5 pg/ml (10.5-21.1)) (U=2091, z=5.70, p<0.001), each compared to controls (6.0 pg/ml (4.7-8.6)) (Fig. 1A-B). The effect size of the sNfL increase was strong both at the preataxic (r=0.54) and ataxic stage (r=0.63). If corrected for the age-related sNfL increase by means of z-transformation, the sNfL increase remained significant in both ataxic subjects (U=1663, z=6.49, p<0.001, Bonferroni-corrected for three groups) and preataxic subjects (U=2208, z=6.54, p<0.001), again with high – and for preataxic subjects even higher – effect sizes (preataxic: r=0.62, ataxic: r=0.63) (Fig. 1C-D). Moreover, sNfL increased from the preataxic to the ataxic stage, as indicated by significant increases of both absolute levels (U=301, z=4.67, p<0.001, r=0.74) and age-corrected levels (U=366, z=2.89, p=0.009, r=0.46). sNfL levels differentiated preataxic and ataxic SCA1 subjects with high accuracy (AUC=0.94 (0.87-1.00), 95% confidence interval).

**Figure 1.**
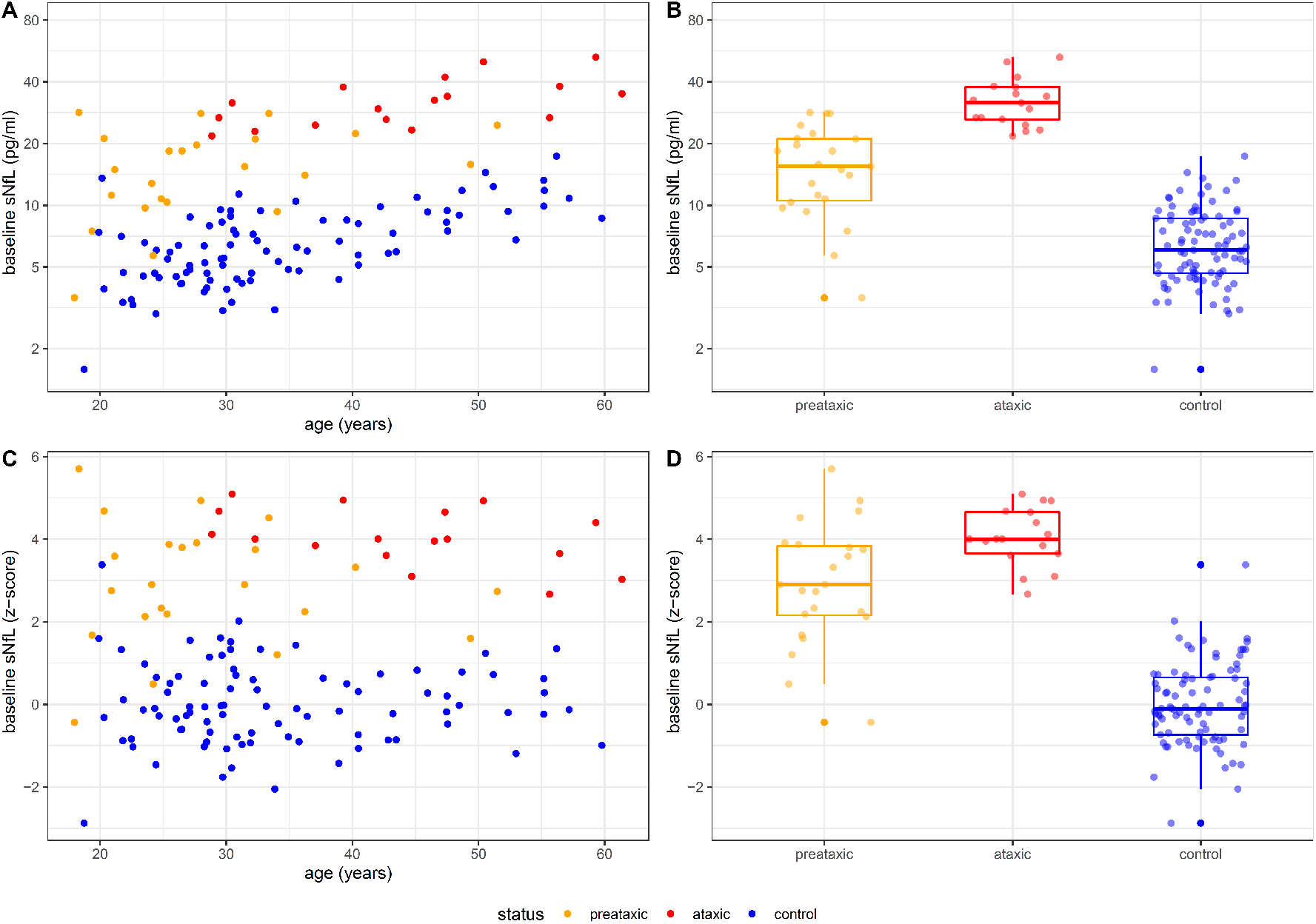
Baseline NfL levels in serum at the preataxic and ataxic stage of SCA1. **(A)** Serum levels of NfL (sNfL) were measured in SCA1 mutation carriers at the preataxic (orange) and ataxic (red) disease stage, and in mutation-negative controls (blue). **(B)** Boxes visualise median, lower, and upper quartiles, whiskers extend to data within 1.5·IQR of the median. **(C)-(D)** To take into consideration the age-related increase of sNfL levels, the absolute levels were corrected for the age-related increase observed in controls by z-transformation.

### Cerebrospinal fluid NfL levels are increased in preataxic and ataxic SCA1, even on the single-subject level

In CSF, NfL levels of each single preataxic (1030 pg/ml) and ataxic SCA1 subject (median: 2971 pg/ml, range: 2164-4032 pg/ml) were clearly above the age-specific normal range of controls (Fig. 2), thus paralleling the NfL increases in blood. If compared to levels of controls of similar age (range: ±5 years), the cNfL increases were significant in both preataxic (V=0, p=0.002) and ataxic (U=150, z=3.51, p<0.001) SCA1 subjects, yielding high effect sizes (preataxic: r=0.48, ataxic: r=0.62).

**Figure 2.**
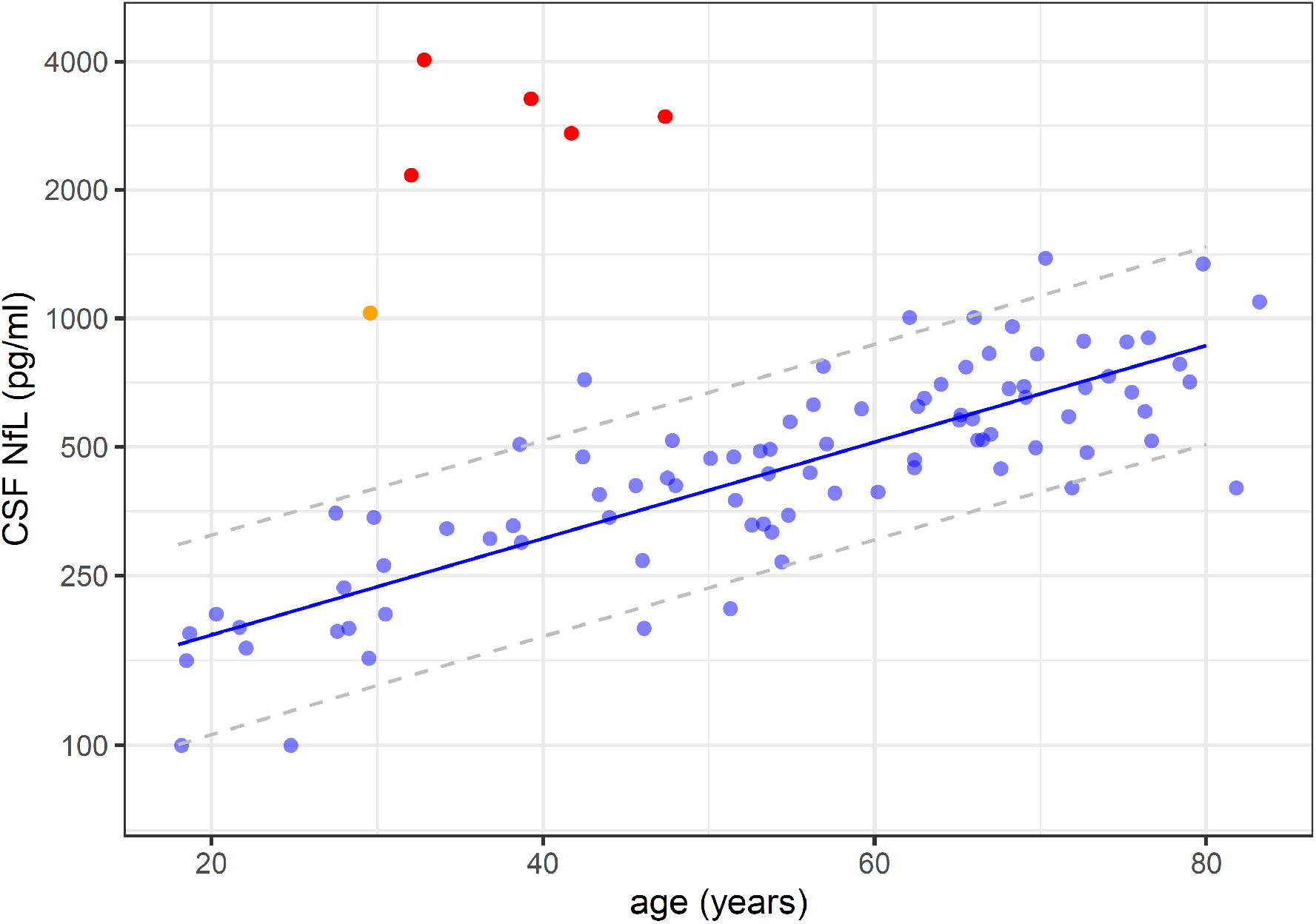
Cerebrospinal fluid NfL levels at the preataxic and ataxic stage of SCA1. NfL levels in the cerebrospinal fluid (CSF) were increased in preataxic (orange) and ataxic SCA1 mutation carriers (red) compared to control subjects (blue). The dashed lines represent the 90% prediction interval of the CSF NfL levels in controls, modelled by linear regression of the log-transformed data.

### Association of serum NfL levels in SCA1 with repeat length, age, and disease stage

We modelled the baseline sNfL levels (log-transformed) across all SCA1 carriers by linear regression with the predictors age and *ATXN1* CAG repeat length, using the pooled data of both preataxic and ataxic subjects (Fig. 3A). Both age (p<0.001) and repeat length (p<0.001) were highly significant predictors of the sNfL level (F (2, 34) = 28.95, p<0.001, adjusted R^2^ = 0.61). The model demonstrated that, for a given age, each increase in the CAG repeat count was associated with higher sNfL levels. For a given CAG repeat count, in turn, the sNfL level increased with age. Additional interaction terms and quadratic terms were not significant. Inclusion of disease stage (i.e., preataxic vs. ataxic stage) as an additional predictor in the model further improved model fit (F (3, 33) = 22.50, p<0.001, adjusted R^2^ = 0.64) and demonstrated that disease stage was a significant predictor of sNfL levels (p=0.049), independent of the other significant predictors age (p<0.001) and repeat length (p<0.001) (Fig. 3B).

**Figure 3.**
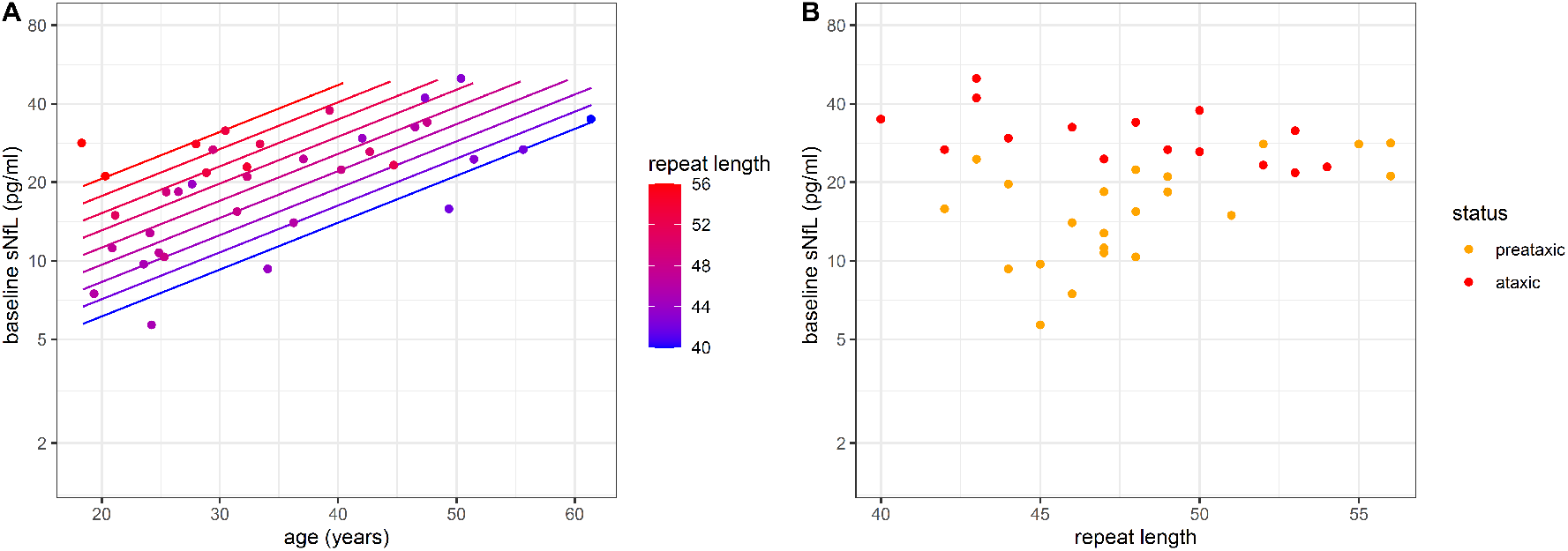
Association between sNfL levels, age and CAG repeat count in SCA1. **(A)** We modelled sNfL levels (log-transformed) across all SCA1 carriers (n=40) by linear regression with the predictors age and *ATXN1* repeat length. Both predictors proved significant and were used to generate the diagram. sNfL levels increased with age and, for a given age, each increase in the repeat length was associated with higher sNfL levels. **(B)** sNfL levels were higher in ataxic (red) than preataxic (orange) SCA1 carriers, also if stratified by *ATXN1* repeat length.

### sNfL levels at the ataxic stage of SCA1 are not associated with disease duration or severity

The sNfL levels in ataxic SCA1 subjects did not correlate with disease duration (ρ=0.38, p=0.169) (Supplement 3A) despite the presence of clinical progression (Supplement 4), supporting the notion that sNfL levels are biomarkers reflecting the rate of ongoing neuronal decay ^11, 22^, where stable increases of sNfL levels might reflect stable progression at the ataxic stage of SCA1 (Fig. 7B). In line with this notion, sNfL levels of ataxic SCA1 subjects also did not correlate with clinical disease severity, as assessed by the Scale for the assessment and rating of ataxia (SARA) (ρ=0.08, p=0.747) (Supplement 3B), suggesting that sNfL levels do not reflect clinical disease severity per se.

### sNfL levels at the preataxic stage of SCA1 increase with proximity to ataxia onset, with significant increases ≥4 years before conversion to the ataxic stage

In preataxic SCA1 subjects, sNfL levels significantly increased with proximity to the individually *predicted* onset of ataxia, as revealed by linear regression (F (1, 20) = 28.67, p<0.001, adjusted R^2^ = 0.57) (Fig. 4A). We calculated the predicted age at ataxia onset based on each subject’s CAG repeat length and their age at blood sampling ^24^. The increase of the sNfL levels with proximity to the *predicted* onset of ataxia remained significant if corrected for subjects’ age at blood sampling (F (2, 19) = 14.63, p<0.001, adjusted R^2^ = 0.56). This finding was confirmed by analysing the actually *observed* onset of ataxia of those preataxic SCA1 subjects for whom conversion to the ataxic disease stage was actually clinically observed during longitudinal follow-up (n=11). Here, sNfL levels significantly increased with proximity to the observed onset (F (1, 9) = 15.53, p = 0.003, adjusted R^2^ = 0.59) (Fig. 4B). This sNfL increase with proximity to the observed onset of ataxia also remained significant if corrected for subjects’ age (F (2, 8) = 6.82, p=0.019, adjusted R^2^ = 0.54). In comparison to controls, the preataxic SCA1 subjects showed significantly increased sNfL levels already 5 years before the predicted ataxia onset and 4 years before the observed ataxia onset, respectively, corresponding to the time point at which the prediction intervals of the sNfL levels in carriers and controls did not overlap any further (Fig. 4A-B).

**Figure 4.**
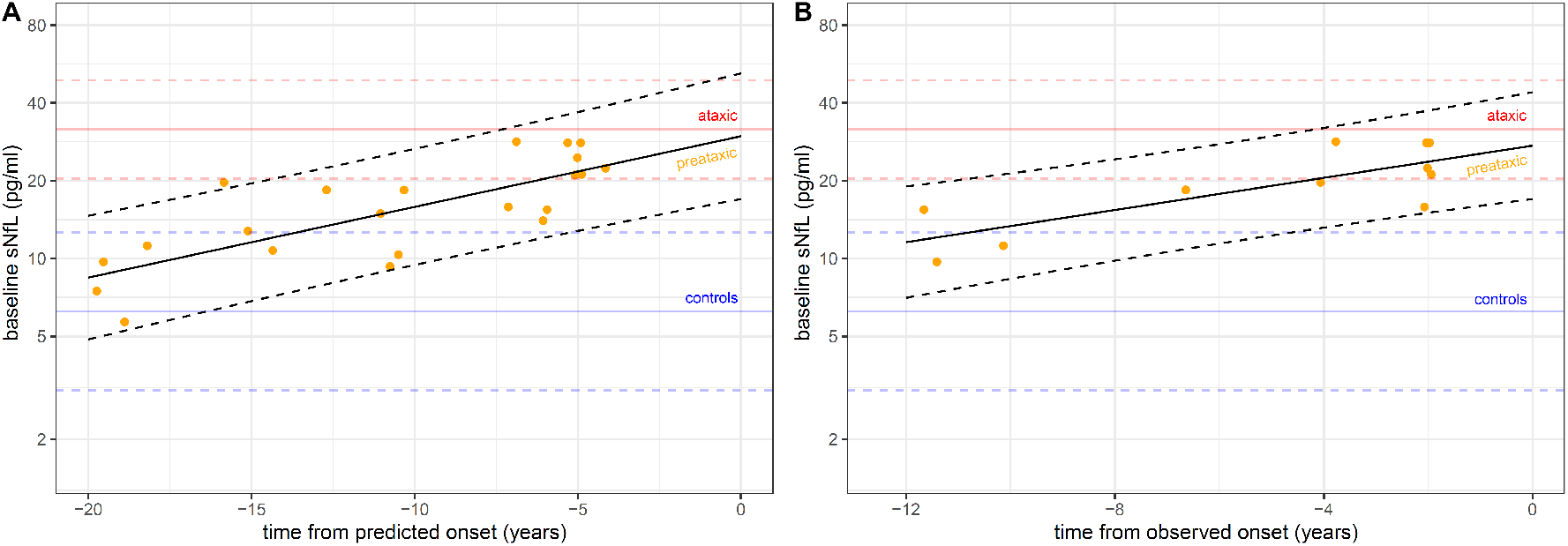
Temporal dynamics of serum NfL levels in preataxic SCA1. **(A)** sNfL baseline levels in preataxic SCA1 subjects were plotted over the time to the individually *predicted* onset of ataxia, with negative time values corresponding to the preataxic disease stage. sNfL levels significantly increased with proximity to the *predicted* onset (F (1, 20) = 28.67, p<0.001, adjusted R^2^ = 0.57), as visualised by the regression line and its 90% prediction band. **(B)** For the subset of preataxic SCA1 subjects for whom conversion to the ataxic disease stage was actually clinically *observed* during follow-up, the sNfL levels at baseline were plotted over the time to the actually observed onset of ataxia. sNfL levels significantly increased with proximity to the observed onset (F (1, 9) = 15.53, p = 0.003, adjusted R^2^ = 0.59). To benchmark the levels of the preataxic subjects (orange points), the levels of controls (blue) and ataxic SCA1 subjects (red) were each visualised as horizontal lines (mean and 90% prediction band).

### Stratifying preataxic SCA1 subjects by sNfL levels regarding the time to ataxia onset

Baseline sNfL levels allowed stratifying preataxic SCA1 subjects with regard to individual time to predicted ataxia onset, as revealed by Kaplan-Meier analyses (Fig. 5A): The median time to predicted ataxia onset was significantly shorter in preataxic subjects with high sNfL levels (5.3 years, sNfL>15.5 pg/ml) than in those with low sNfL levels (12.7 years, sNfL<15.5 pg/ml, threshold for sNfL levels defined by median split) (Χ^2^ (1) = 8.0, p=0.005, log-rank test). Accordingly, for preataxic subjects with high sNfL levels, the hazard of developing ataxia at any given time was approximately 3 times as high as for subjects with low sNfL levels (hazard ratio and 95% confidence interval: 3.58 (1.40-9.11), p=0.008, Cox regression). These findings were confirmed in the subset of preataxic SCA1 subjects for whom longitudinal follow-up information was available (n=18), comprising of 11 converters and 7 non-converters (Fig. 5B). Kaplan-Meier analyses showed that the median time to conversion was significantly shorter in subjects with high sNfL levels (3.8 years, sNfL>17.1 pg/ml pg/ml) than in those with low sNfL levels (11.4 years, sNfL<17.1 pg/ml pg/ml, threshold for sNfL levels defined by median split in the subset of preataxic subjects with follow-up data) (Χ^2^ (1) = 9.8, p= 0.002, log-rank test). For preataxic subjects with high sNfL levels, the hazard of conversion at any given time was approximately 14 times as high as for subjects with low sNfL levels (14.40 (1.71-121), p=0.014, Cox regression). These findings suggest that sNfL levels might be used to stratify preataxic SCA1 carriers with regard to proximity to ataxia onset.

**Figure 5.**
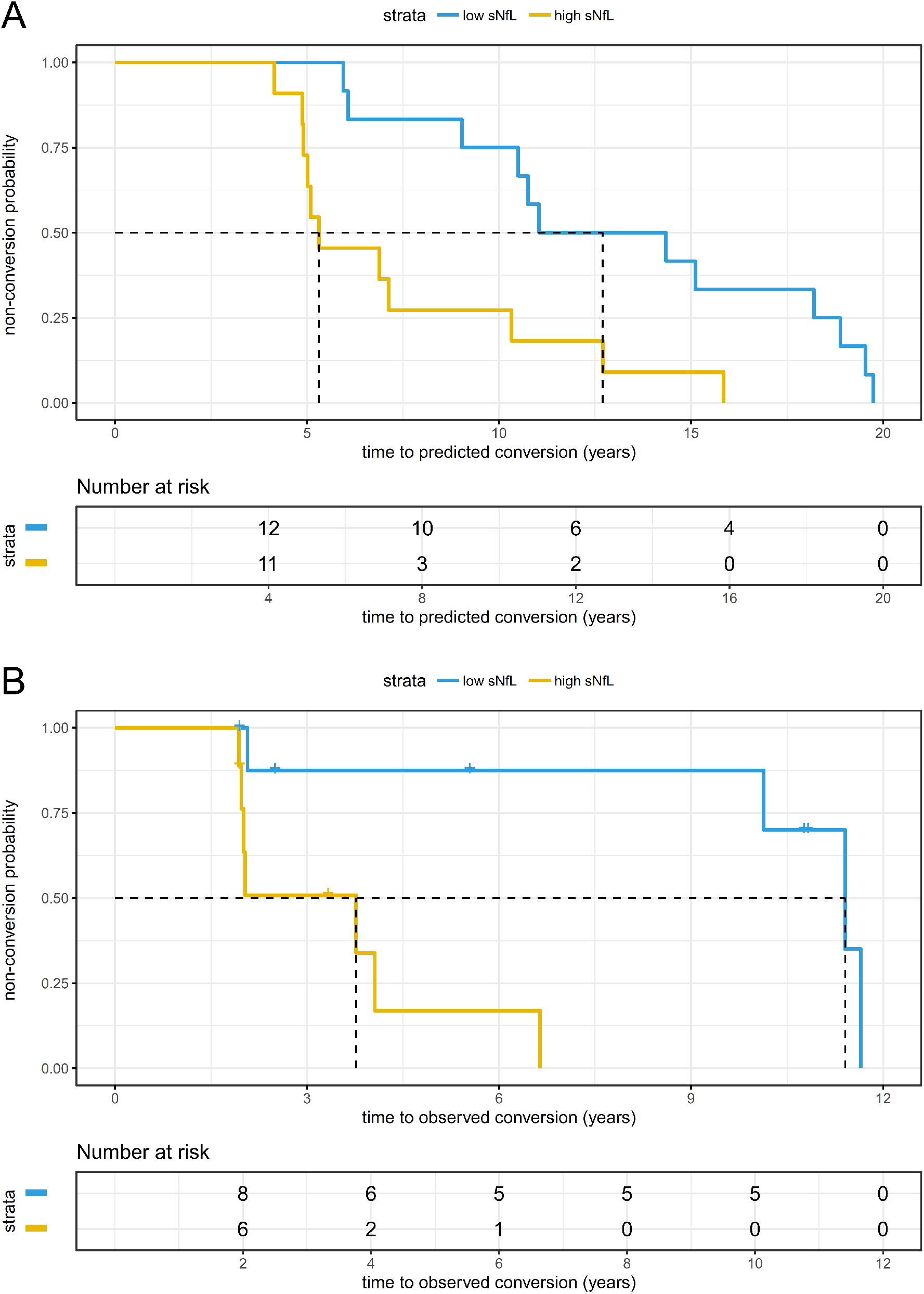
Stratifying the time to conversion to the ataxic disease stage by sNfL levels. **(A)** For all available preataxic SCA1 carriers (n=23), the Kaplan-Meier diagram shows the share of the preataxic carriers remaining preataxic (i.e., the non-conversion probability) plotted over the time to the individually *predicted* ataxia onset, stratified by baseline sNfL level. We calculated the predicted age at ataxia onset based on each subject’s repeat length and their age at blood sampling. We defined high vs. low sNfL levels by median split (threshold: 15.5 pg/ml). **(B)** For the subset of SCA1 carriers for whom longitudinal clinical follow-up information was available (n=18), the diagram shows the non-conversion probability plotted over the time to the *observed* ataxia onset, again stratified by baseline sNfL level (median split, threshold: 17.1 pg/ml). The vertical dashed lines represent the follow-up duration at which 50% of preataxic subjects (in each stratum) have converted to the ataxic disease stage (i.e., median event free survival).

### sNfL levels are increased in preataxic SCA1 subjects without pontine or cerebellar atrophy

In the subset of preataxic SCA1 carriers of whom volumetric MRI data was available (n=12), we identified a substantial subset of preataxic subjects (9/12 subjects, 75%) who did not have any evidence of pontine or cerebellar atrophy (Fig. 6B). sNfL levels of these subjects (16.7 pg/ml (12.4-21.4)) were significantly higher than those of controls (6.0 pg/ml (4.7-8.6)) (U=818, z=4.58, p<0.001, r=0.46), thus suggesting that the sNfL increase in blood might precede not only clinical onset, but also pontine and cerebellar volumetric atrophy, which are considered the earliest and most change-sensitive volumetric MRI changes in SCA1 (Fig. 6A). In the preataxic SCA1 subjects with available MRI data, the sNfL level was not associated with pontine (ρ=-0.07, p=0.834) or cerebellar volume (ρ=-0.03, p=0.921), suggesting that the sNfL elevation in preataxic SCA1 was not driven by the volumetrically measurable brain atrophy (Fig 6C-D), but that neuronal decay might start already before.

**Figure 6.**
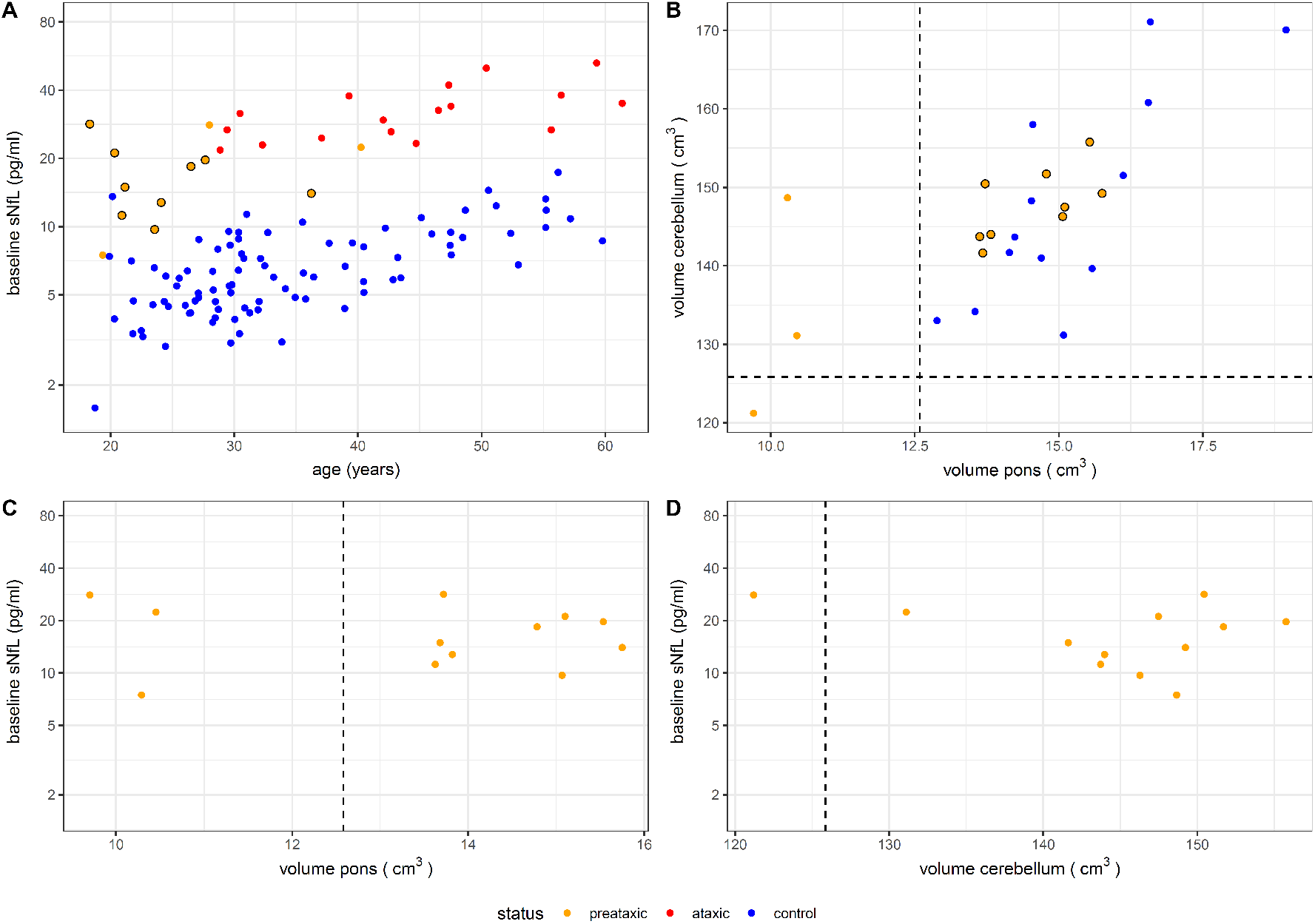
sNfL levels in preataxic SCA1 in relation to pontine and cerebellar volumes. **(A)** sNfL levels of preataxic SCA1 carriers *not* showing any signs of pontine or cerebellar atrophy (orange point *with* black circle) are displayed in comparison to mutation-negative controls (blue), ataxic SCA1 carriers (red), and preataxic SCA1 carriers with pontine or cerebellar atrophy (orange point *without* black circle). **(B)** We considered signs of pontine or cerebellar atrophy present if the respective brain volumes were below a threshold defined as the lower boundary of the 90% confidence interval (i.e., the 5% quantile) of the respective volumes measured in healthy age-matched controls, visualised by dashed lines. The preataxic subjects without signs of pontine or cerebellar atrophy are again visualised by orange points with black circles. **(C)-(D)** In preataxic SCA1 carriers, pontine and cerebellar volume were each not associated with the sNfL level.

### sNfL levels as outcome parameters for treatment trials in SCA1

Our longitudinal measurements demonstrated that intra-individual sNfL levels are stable at the ataxic stage of SCA1 (Fig. 7A-B), with levels not differing between baseline and follow-up visit (t (10) = -0.07, p=0.943, two-sided paired t-test). Based on the longitudinal measurements from baseline to first follow-up (interval: 2.7 years (2.0-3.4)), we estimated sample sizes for future treatment trials using the reduction of sNfL levels as outcome parameter (log-transformed data). Our estimates indicate that, to detect a therapeutic effect size of 50%, the required total sample size would be 14 subjects at the ataxic stage (Fig. 7C, also for visualisation of a range of other possible therapeutic effect sizes).

**Figure 7.**
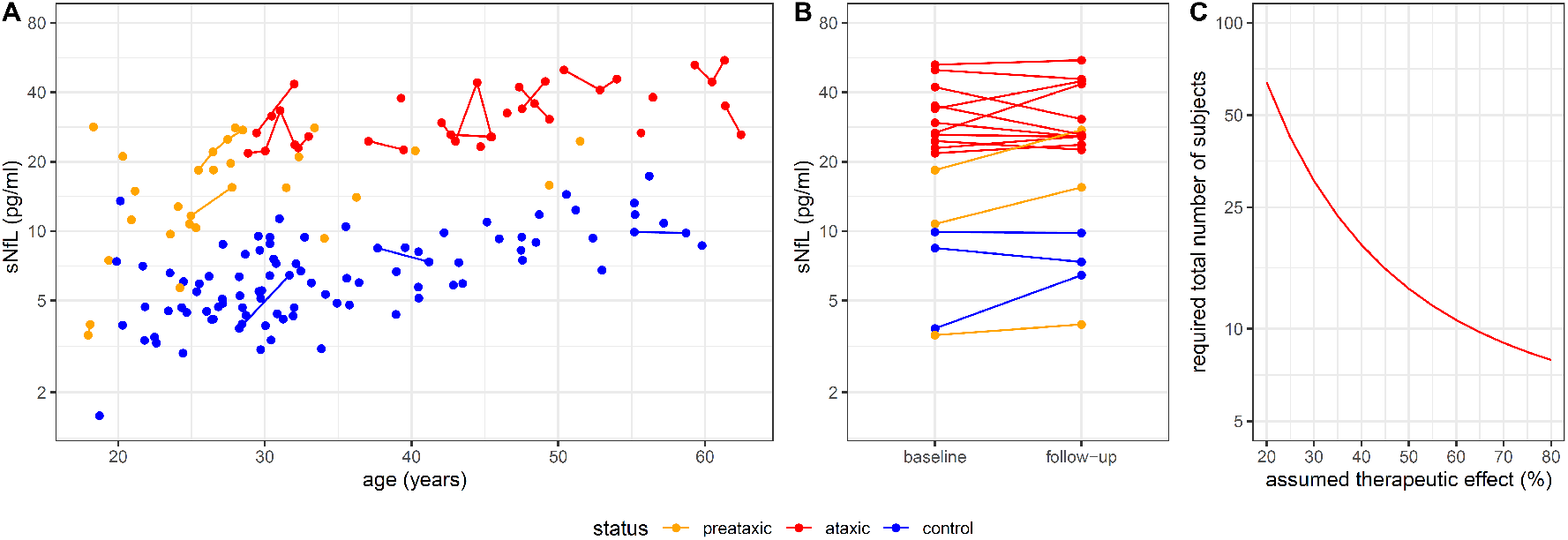
Longitudinal sNfL levels in SCA1 and sample size estimates for treatment trials. **(A)** Longitudinal measurements of sNfL levels were available for a subset of preataxic (orange) and ataxic (red) SCA1 carriers and controls (blue). **(B)** Intraindividual stability of sNfL levels was assessed from baseline to first follow-up visit (interval: 2.7 years (2.0-3.4), median and IQR). Lines link data of the same individuals. **(C)** Sample size estimations were performed for future intervention trials using the reduction of sNfL levels as outcome measure. The estimated total sample size (i.e., sum of subjects in both study arms) was plotted over the assumed therapeutic effect for lowering the neurofilament level in ataxic mutation carriers towards the levels observed in healthy controls.

## Discussion

Leveraging a multicentric cohort of SCA1 mutation carriers, our study demonstrates increased blood levels of NfL in SCA1 (1) not only at the ataxic, but also already at the – likely most treatment-relevant – preataxic stage, (2) paralleled by increased NfL levels in the CSF, (3) rising in proximity to both predicted and actually observed onset, (4) preceding pontine and cerebellar atrophy on volumetric MRI at the preataxic stage, and (5) allowing small sample sizes in future treatment trials.

Our NfL findings from the ataxic stage of SCA1 extend previous findings from monocentric, mixed cohorts of polyglutamine SCAs ^11, 12^ by validating the sNfL increase in SCA1 within a multicentric setting – as essential for clinical trials in rare diseases like SCA1 – and by demonstrating a parallel increase in the CSF, present even on a single-subject level. This parallel increase of NfL levels, which is in line with findings from other neurological diseases ^26, 27^, indicates that peripheral blood levels of NfL indeed offer a valid *peripheral* blood biomarker for degeneration of the *central* nervous system both at the ataxic and preataxic stage of SCA1. However, given the small number of available CSF samples, larger SCA1 cohorts with longitudinal serum-CSF NfL pairs are needed to provide a more fine-grained map of the relative CSF vs. blood NfL dynamics across SCA1 diseases stages, particularly for the preataxic stage.

Moreover, our study suggests that the individual CAG repeat count, age, and disease stage (preataxic vs. ataxic) are the major determinants of sNfL levels in SCA1, thus extending and specifying findings from other CAG repeat-expansion diseases ^22, 23^. The observation that sNfL levels were not closely related to cross-sectional disease duration and disease severity (in line with previous findings ^12, 22^) indicates that sNfL in SCA1 is not primarily a biomarker of the *current state* of disease severity (= disease severity biomarker), but might rather reflect the *rate* of ongoing neuronal decay (= decay rate biomarker) ^11, 22^.

This notion is further supported by the finding that sNfL levels increased from the preataxic to the ataxic stage but did not increase further in the ataxic stage during follow-up despite evidence of clinical progression. Thus, after the increase during the preataxic stage, sNfL levels plateau during the ataxic stage at a stable level. This finding might be best interpreted as evidence that neuronal decay rates increase during the preataxic stage and then remain stably elevated during the ataxic stage. While this notion of stage-dependent NfL dynamics warrants confirmation by larger longitudinal studies, it already receives support from findings in other multisystemic neurodegenerative diseases where NfL levels peak around the time of clinical onset, subsequently stabilise at increased levels, and eventually possibly even decrease in late disease stages ^28, 29^. Our study demonstrates that NfL levels are increased with high effect size not only at the ataxic stage of SCA1, but even already at the preataxic stage – with similarly high effect sizes. Significant increases compared to healthy controls occurred already five years before the predicted ataxia onset, with levels rising further with temporal proximity to ataxia onset. This cross-sectional finding was further supported by intra-individual longitudinal sNfL increases in preataxic subjects. Moreover, the preconversion interval (i.e., the time to predicted ataxia onset) was significantly shorter in the preataxic subjects who had high sNfL levels than those with low levels.

Importantly, these associations of sNfL with proximity to *predicted* ataxia onset were validated by our findings on the associations of sNfL with the *actually observed* ataxia onset: while previous studies in the SCA field mostly relied only on the associations of a biomarker with the *predicted* onset of ataxia – a hypothetical proxy measure based on repeat length and the age reached without developing ataxia ^24^, our study also assessed the associations of sNfL with the actually *observed* onset of ataxia, leveraging longitudinal follow-up data of a sufficiently long time-span to capture the actual disease conversion. These data show that sNfL levels do indeed increase with proximity also to the actual onset, with higher sNfL levels heralding earlier phenoconversion, and with first significant increases already occurring 4 years before the actual ataxia onset.

Taken together, these findings indicate that sNfL levels might serve as biomarkers for stratifying the highly treatment-relevant “proximity-to-onset phase” in SCA1, by allowing stratification of preataxic subjects according to their individual proximity to conversion. A biomarker-based stratification of this phase will be of high relevance for upcoming treatment trials to support the selection of preataxic candidate subjects for intervention trials as potentially burdensome treatments (e.g., intrathecally administered ASOs) should likely be applied neither unnecessarily early nor too late ^3, 22^. However, sNfL might hereby be used not only as a stratification biomarker, but – pending further validation – possibly also as a treatment-response biomarker for the preataxic stage: given the increase of sNfL in the preataxic stage with proximity to clinical disease onset, a treatment-induced reduction of sNfL could serve as an objective biomarker surrogate endpoint for treatment trials at the preataxic stage to capture treatment-induced reduction of the SCA1-related neuronal decay, possibly even on a single-subject level.

sNfL might be useful as biomarker at the preataxic stage of SCA1 also due the fact that it seems to rise rather early during the preataxic stage: it appeared already increased in preataxic subjects with still normal pontine and cerebellar volumes, at least on volumetric MRI. If confirmed in an independent cohort, this finding suggests that the NfL increase is an early event in a multimodal cascade of biomarker alterations in SCA1, preceding even atrophy in those brain regions shown to be most change-sensitive in volumetric MRI of SCA1 ^18, 19^. This notion is in line with recent findings from (murine) SCA3, where it was shown that the onset of sNfL increase already starts with – or even precedes – the onset of Purkinje cell loss ^22^. Similar studies with combined assessment of neuropathology and blood NfL are warranted in SCA1 to further corroborate this hypothesis for SCA1.

Our study provides preliminary sample size estimates for future trials of disease-modifying drugs in SCA1, using the reduction of increased sNfL levels as treatment response biomarker. Our longitudinal assessment of sNfL levels shows that intra-individual biological variation of sNfL levels is likely only a minimal source of noise in observation and intervention trials of SCA1. The use of sNfL levels as outcome parameter might thus help to reduce trial sample sizes in comparison to clinical and other surrogate outcome measures ^14, 18, 19, 30^. For instance, our estimates suggest that a total sample size of 14 subjects would suffice to detect therapeutic effect sizes of 50% at the ataxic stage of SCA1.

Our study has several limitations. Firstly, additional longitudinal measurements of NfL levels during the preataxic stage are needed to model the early intra-individual dynamics of NfL levels in SCA1 in more detail, including non-linear models and sample size estimations at the preataxic stage. Secondly, validation of our findings (in particular of e.g. the MRI findings) is needed in independent larger multicentric cohorts of SCA1 subjects, like the READISCA cohort ^31^.

In conclusion, our multicentric study suggests blood NfL levels as biomarkers for stratification of the preataxic stage (particularly for capturing proximity-to-onset), early neuronal decay at the preataxic stage, and, potentially, treatment response at the preataxic and ataxic in SCA1, and makes first steps towards a multimodal biomarker-based stratification of the preataxic disease stage of SCA1.

## Acknowledgements

We thank all participants and their families for their contribution. CW, LS, TK, AD, RS and MS are members of the European Reference Network for Rare Neurological Diseases Project ID No 739510. This work was supported by the Horizon 2020 research and innovation programme (grant 779257 Solve-RD to MS and RS), the Clinician Scientist Program of the Medical Faculty Tübingen (480-0-0 to CW, and 459-0-0 to DM), the Bundesministerium für Forschung und Bildung (BMBF) through funding for the TreatHSP network (01GM1905 to RS), the European Joint Programme on Rare Diseases (EJP RD) under the EJP RD COFUND-EJP N° 825575 (PROSPAX consortium, to MS, RS and AD, with MS and RS hereby supported by the Deutsche Forschungsgemeinschaft (DFG)). BM was supported by National Scientific Research Programme (NKFI) K 138669 and EFOP 3.6.1-16.2016.00004. The funding sources had no role in the study design, data collection, data analysis, data interpretation, or writing of the manuscript. We thank Elke Stransky for technical assistance and Christoph Kessler for support in setting up the CSF NfL reference cohort.

## Author contributions

CW: design and conceptualisation of the study, subject recruitment, acquisition of data, analysis of the data, drafting and revision of the manuscript.

DM: conduction of laboratory measurements and acquisition of data, revision of the manuscript.

LS, HH, MR, AD, AF, BM: subject recruitment, acquisition of data, revision of the manuscript.

TK: acquisition of funding for the multicentric studies EUROSCA and RISCA, subject recruitment, acquisition of data, revision of the manuscript.

RS: subject recruitment, acquisition of data, analysis of the data, revision of the manuscript.

KR: subject recruitment, acquisition of MRI data, analysis of MRI data, revision of the manuscript

HJ: subject recruitment, acquisition of data, revision of the manuscript.

MS: design and conceptualisation of the study, subject recruitment, acquisition of data, drafting and revision of the manuscript.

## Potential conflicts of interest

CW has nothing to disclose.

DM has served as consultant for Biogen.

LS has nothing to disclose.

HH has nothing to disclose.

MR has nothing to disclose.

TK received consulting fees from Biohaven, Roche, UCB, Uniqure, and Vico Therapeutics.

AD is currently receiving grants from the National Institute of Health (RO1), French National Hospital Clinical Research Program (PHRC), Agence nationale de la recherche (ANR), Triplet Therapeutics, Biogen and Verum, serves on the advisory boards of Roche, Triplet Therapeutics, and Biogen, and holds partly a patent on anaplerotic therapy of Huntington’s disease and other polyglutamine diseases (B 06291873.5).

AF has nothing to disclose.

BM has nothing to disclose.

RS has nothing to disclose.

KR has received grants from the German Federal Ministry of Education and Research (BMBF 01GQ1402, 01DN18022), the German Research Foundation (IRTG 2150), Alzheimer Forschung Initiative e. V. (NL-18002CB), Friedreich’s Ataxia Research Alliance (FARA) and honoraria for presentations or advisory boards from Biogen and Roche.

HJ has nothing to disclose.

MS received consultancy honoraria from Orphazyme Pharmaceuticals and Janssen Pharmaceuticals, both unrelated to the current project and manuscript. He received consultancy honoraria also from Ionis Pharmaceuticals on trial planning in SCA1, yet also unrelated to the current project and manuscript.

## Supplement

**Supplement 1.** Baseline characteristics of the cohort with CSF NfL measurements. Data are reported as median and range. SARA: Scale for the assessment and rating of ataxia.

| group | preataxic carriers | ataxic carriers | controls |
| --- | --- | --- | --- |
| n | 1 | 5 | 89 |
| female sex | 100% | 60% | 48% |
| age (years) | 29.6 | 39.3 (32.1-47.4) | 56.3 (18.2-83.2) |
| SARA score | 0 | 11.5 (4.5-18.0) |  |
| repeat count | 47 | 50 (44-54) |  |
| disease onset (years) |  | 31 (24-39) |  |
| disease duration (years) |  | 7 (3-16) |  |
| cNfL (pg/ml) | 1030 | 2971 (2164-4032) | 474 (100-1384) |

**Supplement 2.**
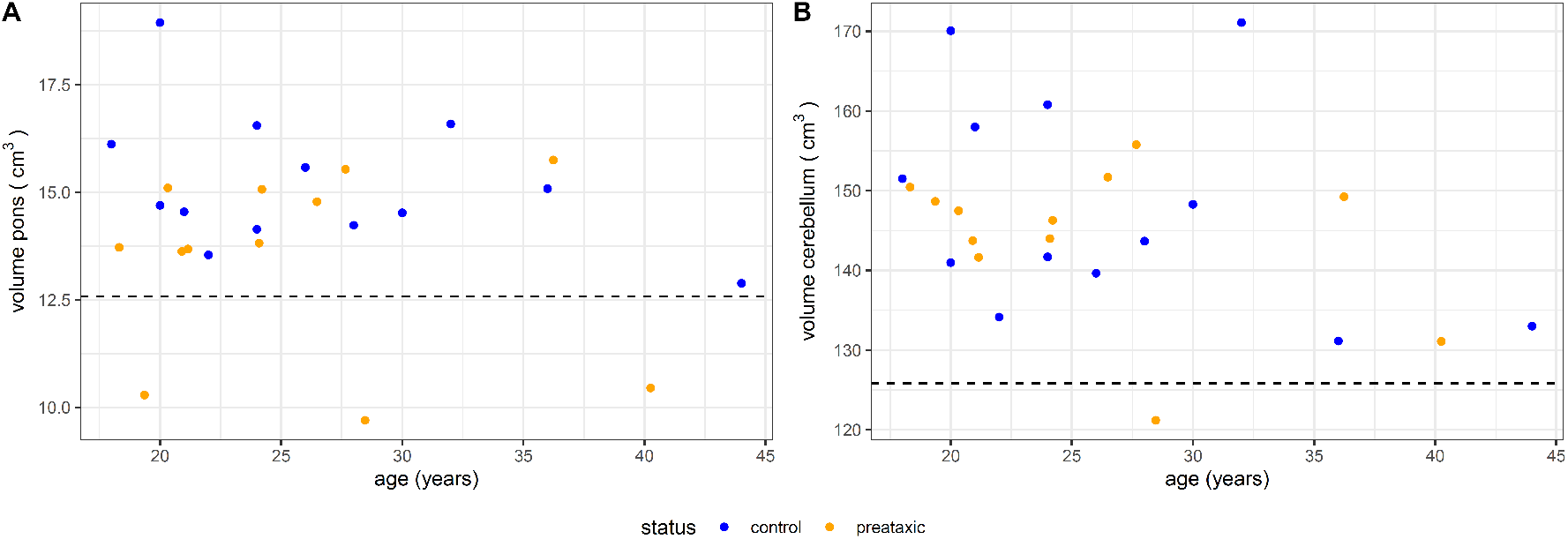
Volumetric MRI of pons and cerebellum in preataxic SCA1 and controls. The volumes of each pons **(A)** and cerebellum **(B)** were assessed by volumetric MRI in SCA1 carriers at the preataxic stage (orange, n=12) and age-matched mutation-negative controls (blue, n=13). We defined the threshold below which we considered the brain volumes of preataxic SCA1 carriers reduced as the lower boundary of the 90% confidence interval (i.e., the 5% quantile) of the respective volumes measured in controls, visualised by the horizontal dashed line. For our further analyses, signs of brain atrophy were considered present if pontine and/or cerebellar volume was reduced below this threshold.

**Supplement 3.**
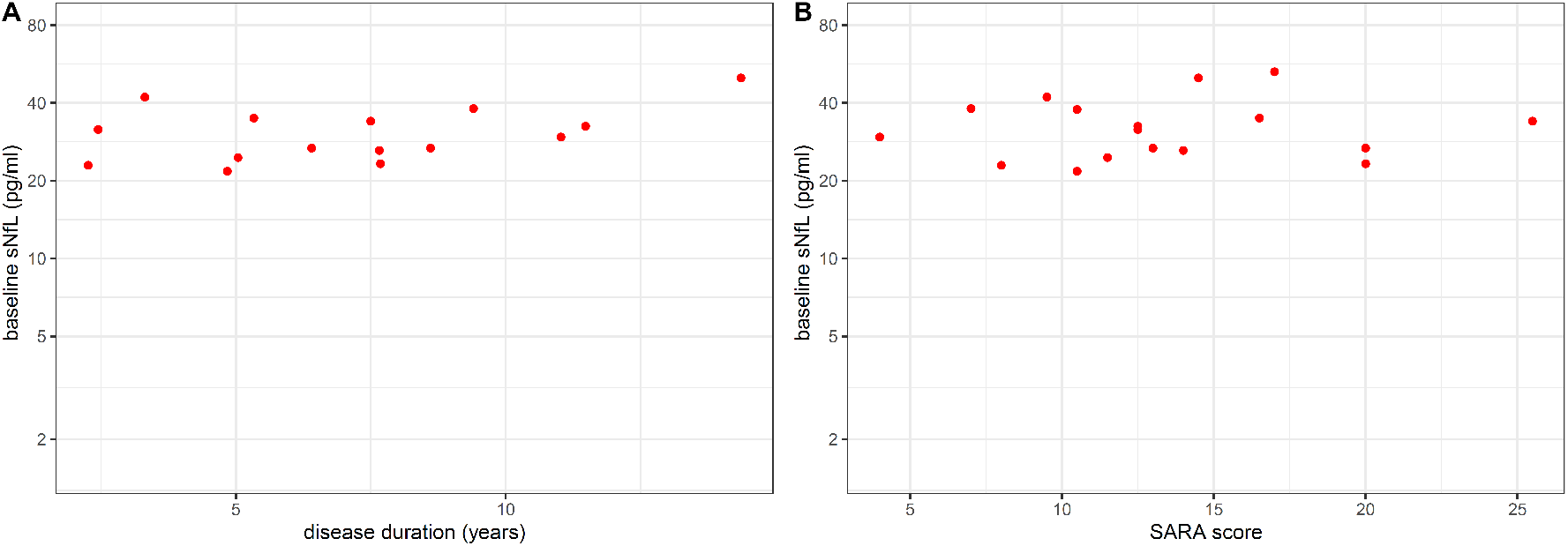
sNfL levels in ataxic SCA1 do not reflect disease duration and severity. **(A)** sNfL levels at the ataxic stage of SCA1 did not correlate with disease duration (ρ=0.38, p=0.169). **(B)** sNfL levels at the ataxic stage of SCA1 also did not correlate with disease severity, as assessed by the Scale for the assessment and rating of ataxia (SARA) (ρ=0.08, p=0.747).

**Supplement 4.**
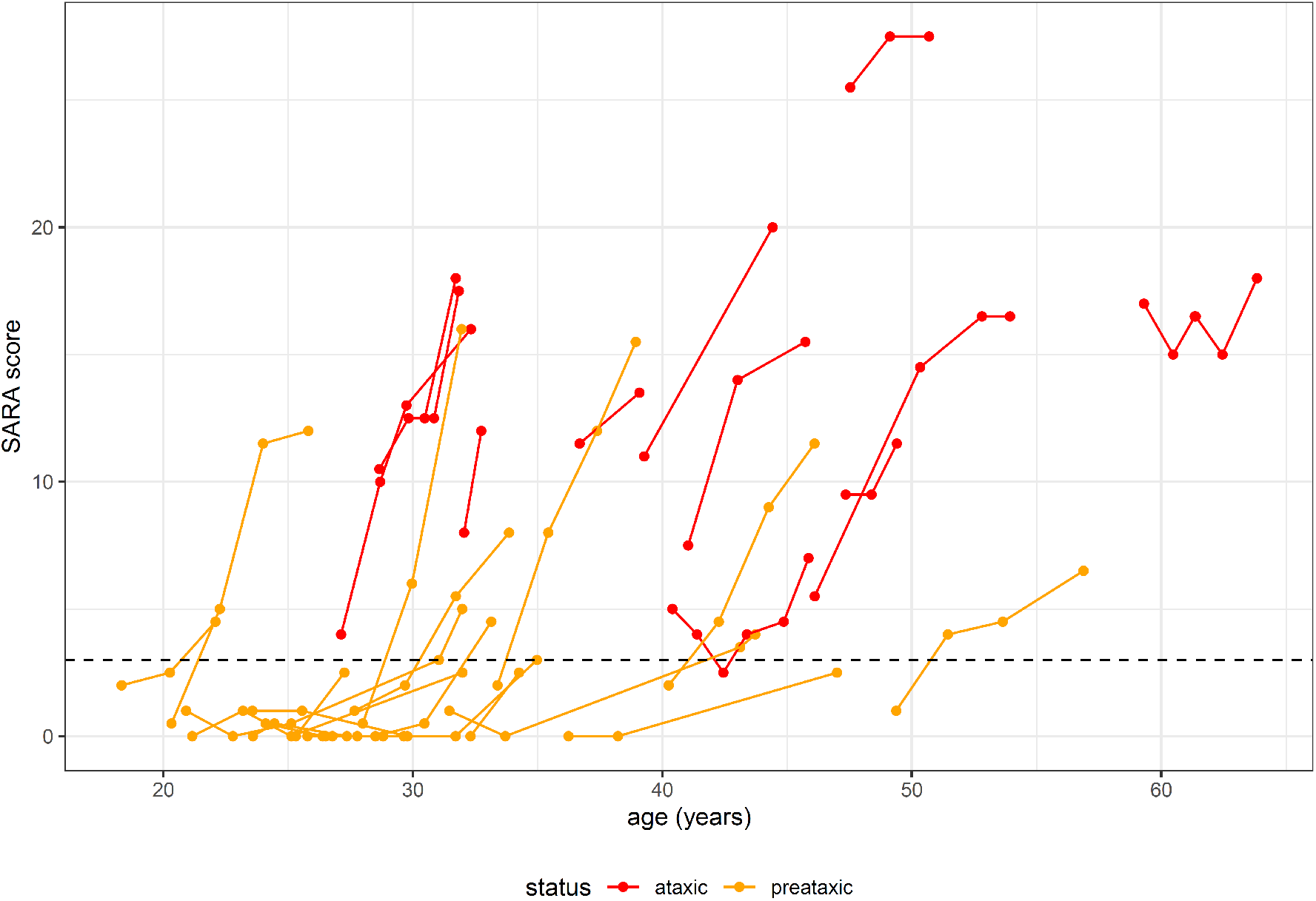
Individual trajectories of clinical severity in preataxic and ataxic SCA1. Disease severity was assessed longitudinally by the Scale for the Assessment and Rating of Ataxia (SARA) in preataxic (orange) and ataxic (red) carriers. Each line connects data of the same individual. The horizontal dashed line visualises the severity threshold (SARA score = 3) below which carriers were classified as preataxic.

## Notes

### Clinical Trial

The EuroSCA cohort was not registered by the respective research consortium. The RiSCA cohort is registered on clinicaltrials.gov (NCT01037777).

### Author Declarations

The ethics committee of the University of Tuebingen approved the study (AZ 103/2017BO2). All subjects provided written informed consent before participation according to the Declaration of Helsinki.

